# Computational link between motivational factors and cognitive deficits in depression

**DOI:** 10.1101/2025.05.30.25328678

**Authors:** Aleks Stolicyn, Liana Romaniuk, Stephen M. Lawrie, Peggy Seriès

**Author notes:** |a*Corresponding author: Aleks Stolicyn Division of Psychiatry, University of Edinburgh Chancellor’s Building, 49 Little France Crescent Edinburgh BioQuarter, Edinburgh EH16 4SB, UK.

## Abstract

**Background:** Cognitive deficits are a common symptom of depression and contribute significantly to the disabling effects of the disorder. Experimentally, they are observed as increased reaction times, increased error rates, and deficient performance adaptation after making errors or receiving adverse feedback, in multiple cognitive paradigms. In the current theoretical study we aimed to address the cause of these cognitive deficits.

**Methods:** We constructed computational models of optimal resource allocation in two cognitive tasks – Delayed Match to Sample (DMS), and Eriksen Flanker (EF). The models explicitly link performance feedback values and beliefs about task controllability with measures of cognitive performance including accuracy, reaction times, and post-error improvement in accuracy (PIA). We then introduced depression-related motivational changes – altered control belief and feedback values (representing learned helplessness, anhedonic valuation and negative bias) – to see if these factors can account for deficits in cognitive performance.

**Results:** In the DMS task, altered control belief and lower valuation of correct performance accounted for decreased accuracy and decreased PIA. In the EF task, altered control belief and lower correct performance valuation could explain increased response times, decreased accuracy and decreased error-related negativity (ERN) signal. Increased valuation of adverse feedback, on the other hand, was linked to increased accuracy and the ERN signal. Furthermore, in the EF task, different combinations of depression-related motivational factors led to different patterns of cognitive performance, which could offer a basis for stratification.

**Conclusions:** Our models offer an explicit computational and algorithmic bridge between the known depression-related motivation factors (learned helplessness, anhedonic valuation) and commonly observed cognitive deficits (increased reaction times, decreased performance accuracy, worse post-error adaptation), which contributes towards a better understanding of depression.

## INTRODUCTION

Depression (major depressive disorder, MDD) is one of the most common psychiatric conditions with an average lifetime prevalence of between 10% and 15%, and 12-month prevalence of around 6% in the general population [1–3]. The two core diagnostic criteria of depression are low mood and loss of interest in daily activities (anhedonia), typically accompanied by other cognitive and somatic symptoms, for a period of at least two weeks [4,5]. Depression has been estimated to cause substantial societal and economic burdens, likely at least in part due to changes in cognition [6–9].

One significant clinical feature of depression is the presence of cognitive deficits. Reduced concentration and attention are both part of the symptomatology [4,5,10,11] and have been measured objectively in a range of different experimental paradigms [12–15]. In addition, pronounced deficits have been observed in the domains of working memory, executive function, and processing speed [12–14,16,17]. For executive function, representative changes are seen at the tasks such as the Wisconsin Card Sort test (WCST), Trails test, Stroop task, as well as planning and executive tests from the Cambridge Neuropsychological Test Automated Battery (CANTAB). Working memory deficits are seen in tasks including pattern recognition and the Delayed Match to Sample (DMS) task. The two core measures which index cognitive performance at many of these tasks are reaction / response times and rates of correct responses (percentages of trials or task stages completed successfully). In depression, reaction times are increased (e.g. Stroop and Eriksen Flanker tasks [13,18–20]), and accuracies are decreased (e.g. DMS and the Tower of London tasks [12,13,15]).

One further behavioural effect representative of cognitive deficits in depression is the diminished post-error improvement in accuracy (PIA). Post-error improvement in accuracy is not universal but is often observed in healthy participants across different cognitive paradigms when improvement in performance is possible [21,22]. In depression, diminished PIA effect has been termed ‘catastrophic response to failure’ and has been seen in multiple experimental paradigms with trial-by-trial feedback, including the DMS and the Tower of London tasks [23–35]. In paradigms without explicit feedback, the diminished PIA effect has been found in some studies [36–38] but not in others [39–41], indicating that post-error performance adaptation deficits might be related to negative feedback.

From the neurophysiological perspective, commission of errors is accompanied by two event-related potentials (measured with electroencephalography) – error-related negativity (ERN) and feedback-related negativity (FRN). Briefly, both ERN and FRN are negatively-going frontocentral signal deflections which occur approximately 80 – 120 ms after commission of errors (ERN), or 250 – 300 ms after presentation of negative feedback (FRN) [42–44]. These signals are considered to represent the neural processes behind cognitive adaptation to errors and feedback [42,45,46]. Current evidence indicates that the FRN signal is decreased in depression, potentially linked to altered reward processing [47–50], although increases have been reported in some studies [49,51–54]. Regarding the ERN signal, the evidence is mixed, with increases or decreases considered to be dependent on specific predominant symptoms such as anhedonia or increased threat sensitivity [50,55–57]. This contrasts with anxiety disorders, where robust increases in ERN have been found [56–60].

From the perspective of motivation – cognitive factors which guide behaviour – depression can be characterised by three core changes: decreased estimates of behavioural control, decreased reward sensitivity, and negatively biased affective cognition. Decreased behavioural control is the core tenet of the *learned helplessness* theory of depression. The theory suggests that when animals or humans are continuously exposed to uncontrollable and undesirable events, they learn to adopt passive coping strategies, characterised by apathy and lack of exploration, considered to represent some depressive symptomatology [61–63]. Decreased reward sensitivity, also interpreted as lower valuation of rewards, is closely related to the symptom of anhedonia and has been shown in multiple computational modelling and other studies of depression [64–66]. Finally, negative affective bias is the propensity to attend to and remember negative material. Negative bias is the core feature of cognitive theories of depression, which suggest that the bias is triggered by adverse environmental circumstances and leads to symptoms such as low mood [67–69]. One interpretation of the negative bias from a computational perspective is in abnormally high valuation of adverse outcomes and, consequently, cues for such outcomes (i.e. negative material) [66,70].

Several theoretical accounts have recently linked motivational factors such as outcome valuation and controllability to cognitive performance. One of these accounts is the Expected Value of Control (EVC) theory, which suggests that cognitive control (effort) is deployed on any task in a way so as to maximise the trade-off between the expected rewards and the costs of effort [71–73]. In the EVC account, cognitive performance directly depends on the amount of cognitive effort deployed, which is in turn based on the estimated probabilities and values of outcomes (e.g. positive and negative feedback), and on cognitive effort costs. An advantage of the EVC theory is that it is a principled normative account of optimal cognitive effort allocation that can be put in precise computational terms. The EVC theory has recently been suggested as a framework capable of causally linking motivational aspects of depression, such as decreased reward sensitivity and perceived lack of behavioural control, to deficits in cognitive control [74].

In the current study we aimed to provide a normative theoretical explanation of depression-related performance differences in two cognitive tasks – DMS and Eriksen Flanker (EF). In depression, accuracy is decreased in the DMS task [12,26,75–81], and some evidence indicates that there is decreased PIA effect [24–26]. In the EF task, reaction times are increased, especially at incongruent trials [19,39,40,82–85], while accuracy is either decreased [19,86], increased [39,87] or unchanged [40,82–85,88–90]. Furthermore, some studies reported increased ERN signal in the task [82,83,87], while others found an opposite effect [19,88,89]. The DMS and EF tasks were chosen because they specifically index two cognitive domains important for everyday function but impaired in depression – working memory and attention, and because substantial experimental evidence of deficits in these tasks already exists (please see above).

We aimed to address a key unresolved issue – what causes these depression-related performance deficits? To date this question has only been addressed partly from a mechanistic neurobiological perspective [11,91,92], but not from a normative one [74]. A distinct advantage of a normative explanation is that it can provide critical insight into how cognitive deficits may arise as a result of the core affective features of depression [74], beyond just brain mechanisms, which in turn can offer a novel direction for identifying treatments that are not solely focused on neurobiology [93]. Within our theoretical investigation, we first constructed new formal computational models, consistent with the EVC theory, that link depression-relevant motivational factors to performance at the DMS and EF tasks. We then simulated depression-related motivational changes – decreased controllability estimate, decreased reward sensitivity, and increased negative valuation – to see if these factors can causally explain the commonly observed performance deficits (decreased accuracy at the DMS task and increased reaction times at the EF task). Finally, we aimed to check if different depression-related motivational factors can lead to different patterns of performance at the EF task, which could explain the discrepant results in the previous studies.

## METHODS AND MODELS

### General Model Framework

Our general framework for cognitive tasks is based on the EVC theory, with an addition of an explicit component to model belief of control. Briefly, at each trial of a cognitive task, we assume that participants combine their knowledge of the task, values of outcomes (i.e. error and correct response feedback), costs of cognitive resources (cognitive effort), and their belief about task controllability, to estimate how much effort is optimal to deploy for completing the trial. Consistently with the EVC theory, optimal amount of effort is the one that maximises the trade-off between the expected outcome value and the costs of expended effort, which we here also term *utility* ( *U* ):

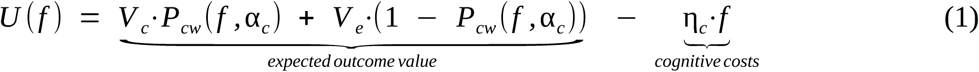

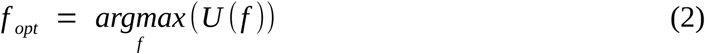

In the above equation *f* represents the amount of cognitive resources deployed at the trial, *V _c_* and *V _e_* are respectively values of correct and error outcomes, η*_c_* is the cost of cognitive effort, and *P_cw_* is the participant’s estimated probability of positive outcome (correct response), which depends on the participant’s knowledge of the task, the deployed cognitive effort and the participant’s belief of control α*_c_*. *f _opt_* is the optimal amount of cognitive effort which maximises utility.

After defining the optimal amount of cognitive resources and completing a single trial, participant receives feedback *F* which they use to update their knowledge of the task or its difficulty. This updated knowledge is subsequently used for defining optimal cognitive resources at the next trial, which serves as the basis for performance adaptation and the PIA effect. Dependent on the cognitive paradigm, these updates can take the form of full statistical learning or a heuristic process of adapting to the task demands drawing on the recent trials.

Given the expected trial outcome value and the actually received outcome *F*, it is possible to compute the valued prediction error signal, which is considered to be reflected in the magnitude of the FRN (ERN) potential in several prominent theoretical accounts [94–97]:

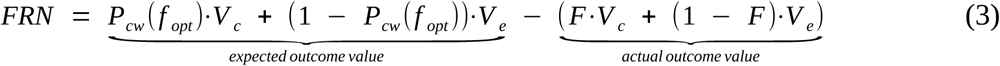

Figure 1 illustrates the general framework within which we further adapted the computational models for the DMS and EF tasks. We below describe our formalisation of these two tasks in more detail.

**Figure 1.**
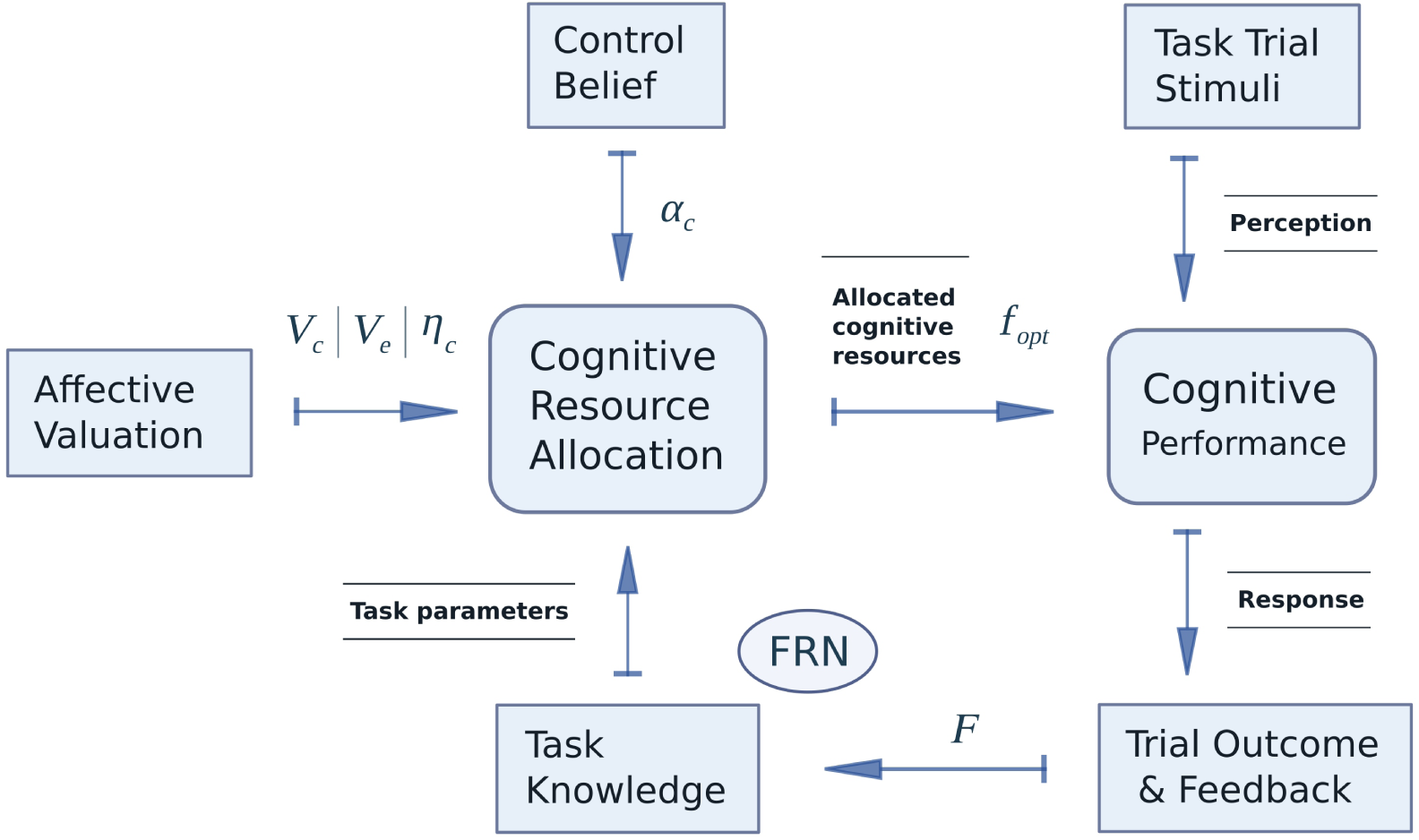
General modelling framework to link motivational factors to cognitive performance. Knowledge of the task, belief of control, and valuation of outcomes and cognitive resources are used to define the optimal amount of cognitive resource to deploy for trial completion. Cognitive resources are then deployed, and trial feedback is received. Feedback is used to update the participant’s knowledge of the task or task difficulty, which are used at the next trial.

### Delayed Match to Sample Model

Briefly, each trial of the DMS task starts with presentation of an abstract visual pattern (the sample), consisting of a number of features. In typical implementations of the task, features are the colours and shapes of the separate quadrants of a rectangular pattern [26,80,98]. The sample then disappears and after a short delay (typically 4 to 12 seconds) a selection of patterns appears (4 patterns in the CANTAB implementation [98]). The participant is tasked with finding and indicating the initial sample pattern in the selection. All patterns in the selection stage are different but share a number of overlapping features. Difficulty of the task is defined by the proportion of overlapping features between the patterns – the more features are the same (overlap) across all patterns, the more difficult it is to identify the original pattern and hence the more difficult the task is. Participant accuracy at finding the original sample, measured across multiple trials, is considered to index working memory capacity [99,100].

We consider that the number of memorised features in the original sample (up to 8 features in our model, consistent with CANTAB) is directly proportional to the amount of the deployed cognitive resources, and is thus subject to the reward-effort based optimisation ( *f* in Equation 1 above). We have analytically derived the correct response probability function dependent on the number of memorised features *f*. We further defined the participant’s belief of control ( α*_c_* ) as their estimate of contingency between actions (the number of memorised features) and outcomes (probability of correct response), consistently with the second notion of behavioural control in Huys & Dayan (2009) [101]. We then applied this definition of control to obtain the function of correct response probability weighted by the control belief ( *P_cw_* ( *f,* α*_c_* ) in Equation 1) – this represents participant’s incorporation of their belief of control in estimation of correct response probability. Figure 2 illustrates the DMS task and how outcome valuation and control belief fit into performance. The effect of belief of control ( α*_c_* ) on the estimated correct response probability ( *P_cw_* ) is illustrated in Figure 3C. As can be seen in Figure 3C, higher belief of control leads to higher estimated contingency between the number of memorised features ( *f* ) and the probability of correct response ( *P_cw_* ) – i.e. each extra memorised feature leads to a steeper increase in the estimated correct response probability. This estimated contingency in turn affects performance by influencing the number of memorised features considered to be optimal for the reward-effort trade-off (Equations 1-2). Task difficulty (i.e. the proportion of overlapping pattern features) was considered to be unknown to the participant, and re-estimated with feedback after each trial. The effect of the DMS task difficulty on the correct probability function, and consequently on the utility function (Equation 1) is illustrated in Figures 3A and 3B. As can be noted in Figure 3B, higher task difficulty leads to higher optimal number of features to be memorised. After errors, participants re-estimate the task difficulty to be higher, and consequently memorise more features in the following trials. This serves as the basis for post-error performance adaptation and the PIA effect. Please see supplementary section S1 for a detailed mathematical description of the DMS task model.

**Figure 2.**
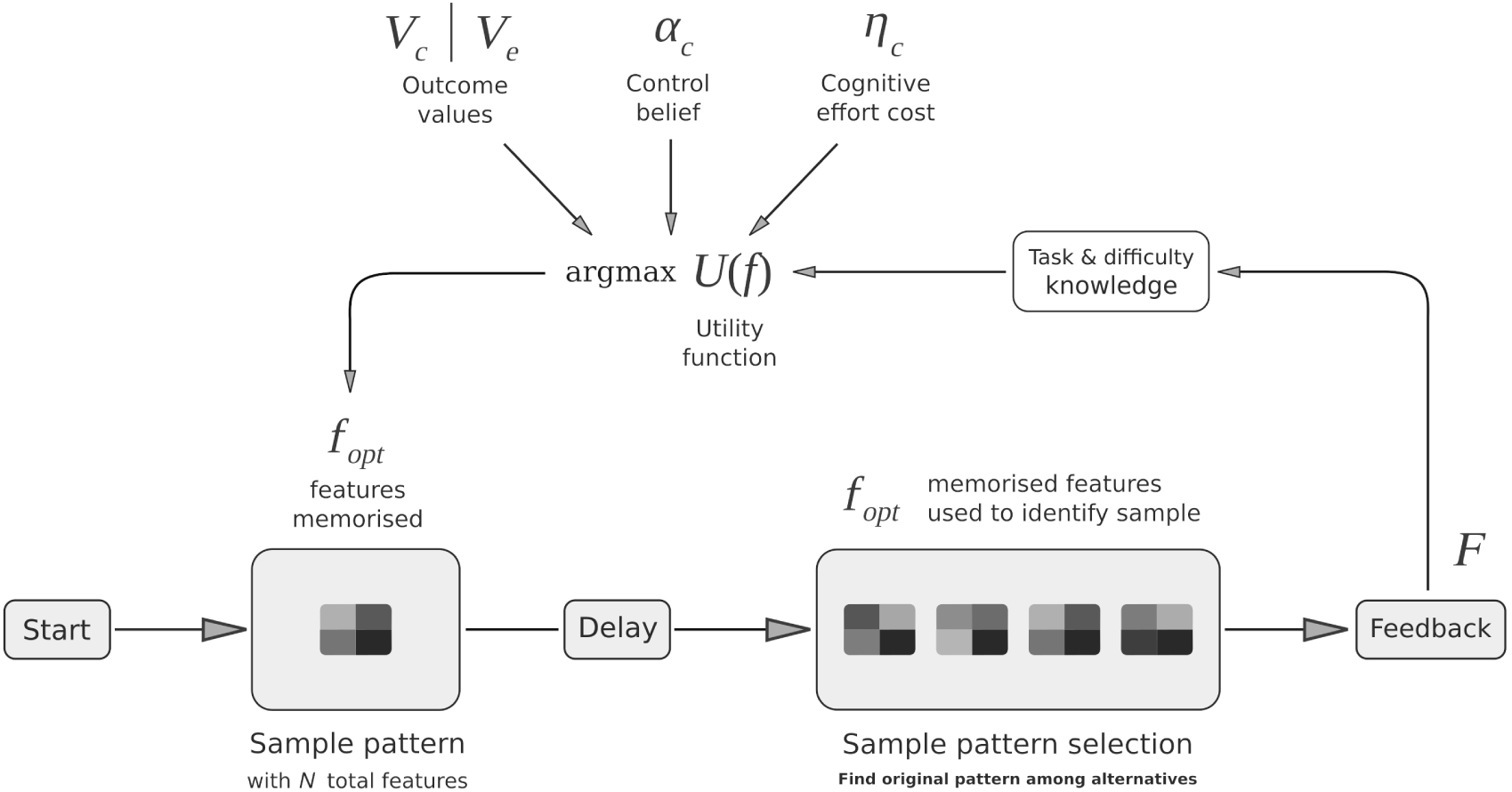
Delayed Match-to-Sample task illustration. Participant is asked to memorise a sample pattern and then to recognise that pattern in a selection after a short delay. They use their knowledge of the task, values of correct and error outcomes ( *V _c_* and *V _e_* ), belief of control ( α*_c_* ), and cost of cognitive effort ( η*_c_* ) to estimate the number of pattern features *f _opt_* to memorise for the best reward-effort trade-off (highest utility *U* ). They then implicitly memorise *f _opt_* features and use them to find a matching sample at the selection stage. *f _opt_* directly affects the probability of correct response. The received trial feedback *F* (correct or error) is then used to update the estimate of the task difficulty, which is used to re-estimate *f _opt_* for the subsequent trial. *Note*: Patterns are for illustration only and do not represent actual stimuli typically used at the task.

**Figure 3.**
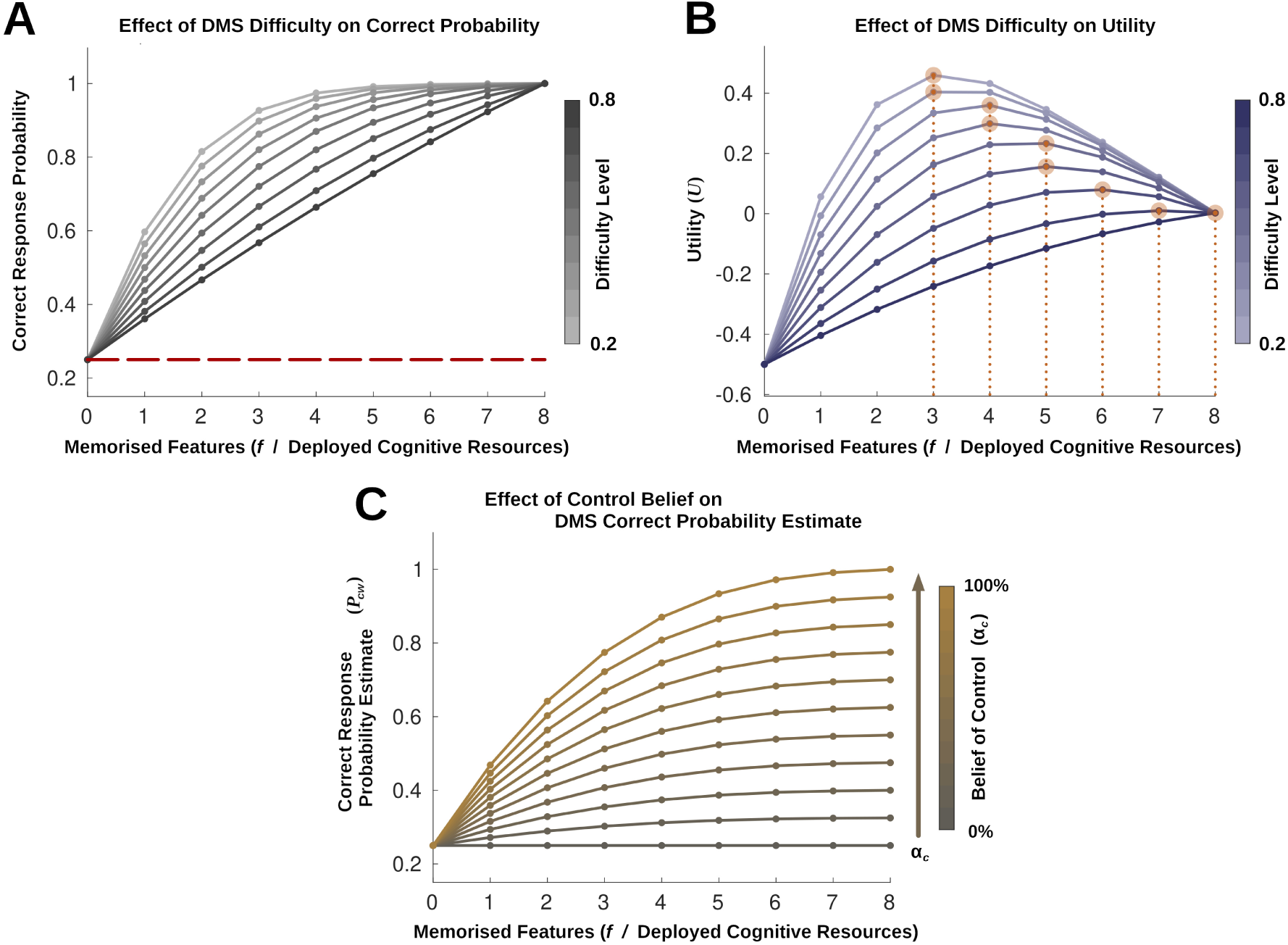
**(A)** Effect of the DMS task difficulty on correct response probability. Difficulty level is defined by the proportion of features which overlap (are the same) across all patterns at the selection stage – the higher the proportion, the more features the participant needs to memorise to correctly identify the original sample. When participant does not memorise any features, they perform at chance level (denoted with the red dashed line). **(B)** Effect of the DMS task difficulty on optimal utility. Orange circles with dotted lines indicate optimal utility (highest *U*, *y* axis) for different levels of difficulty (blue shades), and the corresponding numbers of features to be memorised ( *f*, *x* axis). Higher task difficulty (darker line shades) leads to higher optimal number of memorised features. After errors, participants re-estimate task difficulty to be higher, and consequently memorise more features at the next trials; this leads to post-error improvement in accuracy (PIA effect). **(C)** Effect of the belief of control on participant’s correct response probability estimate. Higher belief of control ( α*_c_*, coloured grey to orange) leads to higher contingency between the number of memorised features ( *f*, *x* axis) and the estimated probability of correct response ( *P_cw_*, *y* axis) – i.e. higher numbers of memorised features are estimated to lead to better chances of success (correct response).

### Eriksen Flanker Model

EF task is a two-alternative forced-choice perceptual decision-making task. At each trial, the participant is presented with a target stimulus which defines the correct response (e.g. arrows ‘<’ or ‘>’ in the arrow version of the task, indicating respectively left or right button presses). The target stimulus is presented with distractors at each side (flankers), which are either congruent with the target and indicate the same response, or incongruent and indicate a conflicting response [102]. In the arrow version of the task, a congruent trial consists of five arrows with the same direction (e.g. ’< < < < <’). Incongruent trials have target and flanker stimuli aimed in different directions (e.g. ’> > < > >’). Difficulty of the task is defined by the proportion of incongruent trials – the more incongruent trials there are, the more attention the participant needs to pay to the target stimulus to remain accurate. Reaction times and error rates at the task are considered to index selective attention and inhibitory cognitive control.

To model reaction times and error rates at the task we applied the drift-diffusion framework of Dillon et al. (2015) [39], which was in turn adapted from Noorani & Carpenter (2013) [103]. This framework is illustrated in Figure 4 and further details are provided in supplementary section S2.1. Briefly, the framework consists of three drift-diffusion processes: prepotent process which defines response times at congruent trials, inhibitory process which prevents errors at incongruent trials, and executive process which defines response times at incongruent trials. Inhibitory and executive processes are related to goal-driven cognitive control and are effortful – the more cognitive resources are allocated to these processes, the faster and more accurate is task performance. We consider that cognitive effort allocated at the EF task is directly proportional to the drift rates of the inhibitory and executive processes ( *v_inh_* and *v_exec_* in Figure 4), that these drift rates incur costs ( η*_c_* in Equation 1), and are subject to the reward-effort based optimisation similarly to the number of memorised features in the DMS task (i.e. equivalent to *f* in Equation 1). We analytically derived the correct response probability function dependent on the inhibitory drift rate, and applied our definition of behavioural control ( α*_c_* ) to obtain the function of correct response probability weighted by the control belief ( *P_cw_* ( *f,* α*_c_* ) ), similarly to the DMS task. Please see supplementary section S2 for a detailed description of our model of the EF task.

**Figure 4.**
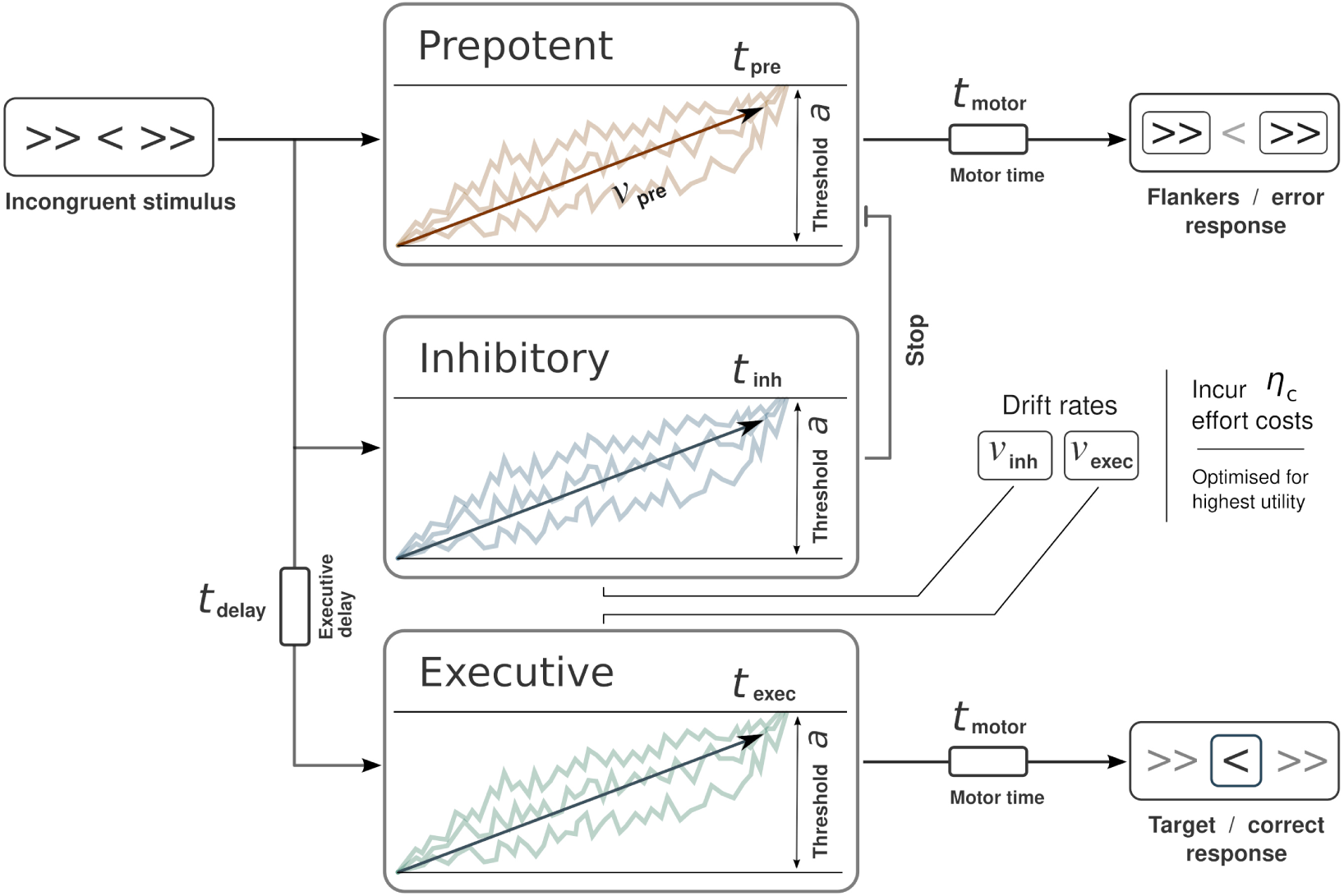
Eriksen Flanker task drift-diffusion model framework [39,103]. *Prepotent* process leads to error responses at incongruent trials. When the *prepotent* process is stopped by the *inhibitory control* process, *executive control* process is able to complete a correct response. Inhibitory and executive processes are effortful and their respective drift rates ( *v_inh_* and *v_exec_* ) incur the cognitive effort costs η*_c_*. These two drift rates are optimised to maximise the reward-effort trade-off (similarly to the number of features *f* in the DMS task).

### Depression Modelling

The DMS and EF models were first specified and parametrised to replicate performance of healthy control participants. We then investigated if introduction of depression-related motivational factors can lead to the cognitive performance deficits. Within the DMS model, we aimed to find affective parameter alterations which can result in decreased accuracy and decreased or eliminated PIA effect. Within the EF model, we aimed to replicate increased correct response times at incongruent trials [39], and to test which parameter changes might lead to decreased or increased accuracies, as well as decreased or increased ERN magnitudes. Following the aims of the study, we constrained investigations of depression to the following three possibilities: (1) a decrease in the α*_c_* parameter (control belief deficit, representing *learned helplessness*); (2) a decrease in the *V _c_* parameter (representing anhedonic decrease in reward sensitivity); (3) an increase in the magnitude of the *V _e_* parameter (representing biased negative valuation, note that *V _e_* is always negative).

## RESULTS

### Delayed Match to Sample Modelling Results

We specified parameters of the DMS model to be broadly consistent with the CANTAB implementation of the task, and set valuation and control parameters of healthy controls to replicate a 90% accuracy [24,26] and a 5% PIA effect. Difficulty of the task (i.e. the proportion of features overlapping between patterns) was set to the value of 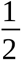. Model performance was obtained by simulating 200 experimental participants with 1000 trials for each participant. Please see supplementary section S1.8 for the exact details of the DMS model parameter constraints and specification.

Simulation results revealed that two out of the three investigated depression-related deficits could replicate decreased accuracy and decreased PIA effect within the DMS model. Performance deficits were replicated by either the decreased control estimate ( α*_c_* ) or the decreased reward value ( *V _c_* ), but not by an increase in the negative outcome value ( *V _e_* ). To compare the two models of depression between each other, we constrained them to produce an approximate 10% decrease in accuracy – this corresponded to a 79% decrease in reward value and a 40% decrease in control belief (supplementary section S1.9). Results of the model simulations can be seen in Table 1 and Figure 5. Simulations predicted a higher decrease in the FRN for the anhedonic deficit as compared to the control belief deficit ( *p* < 0.00001 ), with otherwise similar decreases in accuracy and the PIA effect ( *p* > 0.10 ).

**Figure 5.**
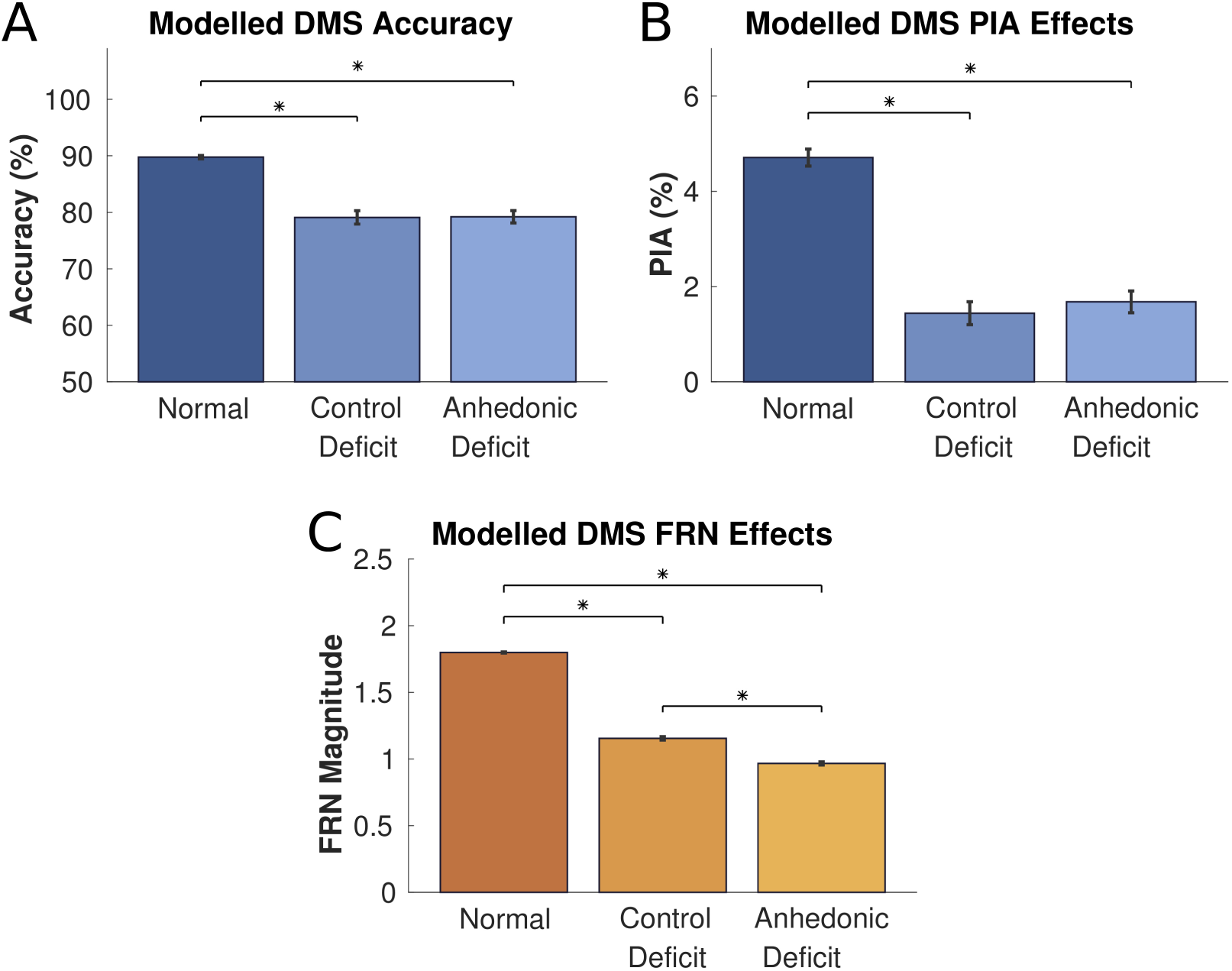
Modelled depression effects over DMS measures: **(A)** Performance accuracy; **(B)** Post-error accuracy improvement (PIA) effect; **(C)** FRN magnitude. Stars indicate significant differences (*p* < 0.00001). Error bars in plots A and C represent standard deviations of the mean. Error bars in plot B represent standard errors of the mean. Control belief and anhedonic valuation deficits result in similar decreases in accuracy and the PIA effect (A and B) but have different effects on the FRN magnitude (C).

**Table 1.** Delayed Match to Sample modelled control and depression measures.

|  | *Accuracy* | *Post-Error*<br>*Error Rate* | *Post-Correct*<br>*Error Rate* | *PIA* | *FRN* |
| --- | --- | --- | --- | --- | --- |
| Control | 89.77% | 6% | 10.71% | 4.71% | 1.80 |
| Reward<br>Sensitivity<br>Deficit | 79.23% | 19.41% | 21.09% | 1.68% | 0.97 |
| Control<br>Belief<br>Deficit | 79.11% | 19.72% | 21.16% | 1.44% | 1.15 |

*Note:* FRN is in abstract units related to the valuation of correct and error performance (Equation 3).

### Eriksen Flanker Modelling Results

Parameters of the EF drift-diffusion model (Figure 4) were specified to closely approximate the values derived for control participants in Dillon et al. (2015) [39], with the exception for the inhibitory and executive drift rates because these are considered to be optimised to maximise the reward-effort trade-off. Difficulty of the task (proportion of incongruent trials) was set to 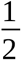. Similarly to the DMS model, performance measures were obtained by simulating 200 experimental participants with 1000 trials for each participant. Please see supplementary section S2.7 for the exact details of the RD model parameter specification. Table 2 summarises the modelled EF performance measures compared to the experimental results of Dillon et al. (2015) [39].

**Table 2.** Eriksen Flanker task control model performance.

|  | **Experimental** | **Modelled** |
| --- | --- | --- |
| *Congruent RT* | 360 ms | 367 ms |
| *Incongruent*<br>*Correct RT* | 448 ms | 448.5 ms |
| *Incongruent*<br>*Error RT* | 319 ms | 328.4 ms |
| *Incongruent*<br>*Correct Rate* | 76.2% | 73.3% |
| *PIA* | 1% | 5.07% |
| *ERN* | - | 1.064 |

*Note:* ERN is in abstract units related to the valuation of correct and error performance (Equation 3).

Simulation results showed that all three depression-related deficits led to increased incongruent correct response times. To compare the three depression models, we constrained them to approximate the mean incongruent correct response time of depressed participants in Dillon et al. (2015) (approx. 475 ms in depression vs. approx. 450 ms control) [39]. To investigate if the different depression-related factors might have counteracting effects when they are present together, we simulated two additional combinations – control belief deficit together with negative value deficit, and reward sensitivity deficit with negative value deficit. Table 3 illustrates qualitative effects of the depression models over task performance (please see supplementary section S2.8 for the exact RD depression model parameter values and the exact effects of depression models over performance). Control belief and reward sensitivity deficits resulted in similar qualitative patterns – increased RT for incongruent trials (both error and correct), decreased incongruent trial accuracy, and decreased ERN magnitude. No change in the PIA effect was observed with these models. The aversive deficit model resulted in an opposite pattern for all measures except the incongruent correct RT. The two additional combined models resulted in spared performance on all measures except for the incongruent correct RT and the ERN, both of which were increased. This suggests that when some of the motivational deficits are present together they may have counteracting effects on cognitive performance.

**Table 3.** Qualitative depression model effects over EF task performance.

|  | *Incongruent*<br>*Correct RT* | *Incongruent*<br>*Error RT* | *Incongruent*<br>*Correct Rate* | *PIA* | *ERN* |
| --- | --- | --- | --- | --- | --- |
| Control Belief Deficit | ↑↑ | ↑ | ↓↓ |  | ↓↓ |
| Reward Sensitivity Deficit | ↑↑ | ↑ | ↓ |  | ↓↓ |
| Negative Value Deficit | ↑ | ↓ | ↑↑ | ↓ | ↑↑↑ |
| Control Belief & Negative Value Deficits | ↑↑↑ |  |  |  | ↑↑ |
| Reward Sensitivity & Negative Value Deficits | ↑↑ |  |  |  | ↑ |

*Note:* Similar qualitative performance patterns are highlighted with similar colours.

## DISCUSSION

### Summary of Results

We constructed two computational models which explicitly link motivational factors (outcome valuation, belief of control) to cognitive performance at the DMS and EF tasks. The models are based on the EVC theory, which posits that the brain allocates cognitive resources in a way so as to maximise reward-effort trade-off [71–73]. We further introduced depression-related motivational factors (anhedonic reward sensitivity, negative valuation and perceived lack of control) in the models to see if they can account for the performance deficits typically seen in depression. Results revealed that lower belief of control (“learned helplessness”) and anhedonic reward sensitivity could account for reduced accuracy, PIA effect and FRN potential at the DMS task, with reduced reward sensitivity leading to higher FRN decrease. At the EF task, different depression-related motivational factors led to different performance patterns, but all resulted in increased correct response times at incongruent trials (Table 3). Overall, the results show that depression-related motivational factors can causally account for cognitive performance deficits, and that patterns of cognitive performance could potentially serve as a basis for stratifying depression according to motivational changes.

### Depression Modelling

Depression-related cognitive deficits within our models arise because of changes in utility estimation. Specifically, altered control belief and outcome valuation lead to higher utility being assigned to the deployment of lower amounts of cognitive resources, and subsequently to poorer performance due to less resources deployed (Equations 1 and 2, Figure 1). As such, our models only provide a normative (rational) explanation of the aetiology of cognitive deficits, but do not consider that they may arise as a consequence of purely mechanistic changes in the brain, which is possible in more severe depression subtypes [11,92,93]. One additional normative explanation of cognitive performance deficits could be in an increase in the costs of cognitive effort ( η*_c_* in Equation 1). Although this is a generally plausible motivational change [74], we did not explore it in this study because there is currently very limited evidence for changes in costs of effort in depression [104,105], in contrast to the changes in outcome valuation and control belief [61–66,70].

### Modelling Predictions

Apart from causally linking motivational factors to cognitive performance, our model of the DMS task made a specific prediction about the magnitude of the FRN signal [94–97]. Specifically, the anhedonic reward sensitivity model of depression predicted a larger decrease of the FRN signal compared to the control belief deficit, with otherwise comparable decreases in accuracy and the PIA effect (Figure 5). This prediction could be directly tested experimentally by evaluating a group of depressed participants with higher anhedonia scores and lower helplessness scores against a group with higher helplessness and lower anhedonia scores. The former group should display lower FRN magnitudes compared to the latter, when both groups are matched for the DMS task accuracy.

Regarding the EF task, our simulations revealed three distinct qualitative patterns of depression-related performance (Table 3), which supports the notion that depression may be stratified on the basis of performance at cognitive tasks. Decreased accuracy and decreased ERN signal were attributed to lower control belief or reward sensitivity and could thus be indicative of responsiveness to behavioural activation therapy [106], or to dopaminergic medication [107,108]. Increased accuracy and increased ERN signal, on the other hand, were related to increased negative valuation and could thus indicate responsiveness to anxiolytic treatments such as higher doses of SSRI or cognitive behavioural therapy [109–111]. Relation of cognitive performance to optimal depression treatment selection remains an interesting and important avenue for future work.

### Neurobiological Interpretation

As regards the neurobiological interpretation, the core resource allocation computation (Equations 1-2) is theorised to be completed in the dorsal anterior cingulate cortex (ACC) within the EVC account, which is supported by a range of human and animal studies [71–73]. Outcome values could be extracted from long-term affective memory in the amygdala and the ventral striatum upon first presentations of positive and negative feedback [112,113], then maintained in the orbitofrontal cortex and the insula [114–117], and signalled to the ACC for resource allocation computations [71,72,117]. Belief of control is likely maintained and signalled by multiple areas of the prefrontal cortex, but also possibly by tonic levels of serotonin [61–63,118]. Actual control over task execution is likely performed by specific sub-regions of the lateral prefrontal cortex (LPFC), dependent on the requirements of the task [73,119]. Prime candidates for cognitive resources are the neurotransmitters needed to modulate neural activity in the LPFC, as well as the sources of metabolic energy for prolonged firing of neural cells. These neurotransmitters include dopamine, noradrenaline and acetylcholine [120–123], whereas the known depletable metabolic energy sources in the brain include blood glucose, adenosine triphosphate and astrocytic glycogen [73,124–126]. How exactly the ACC may direct these neural resources for task completion remains an interesting avenue for future work.

### Limitations

Despite the explanatory and predictive advantages of our computational account, there are some limitations. First, we modelled optimal cognitive resource allocation in terms of explicit computations of outcome probabilities and utility (Equations 1-2, supplementary sections S1.2-S1.4 and S2.2-S2.4). In practice, however, it is more likely that outcome probabilities ( *P_cw_* in Equation 1) are either approximated or gradually learned over time and cached in the brain, to reduce computational complexity. This process could take place over the initial trials of the task and engage neural systems for reinforcement learning [127]. To focus on the effects of interest, we did not model complete statistical or reinforcement learning and instead constructed a simplified account of continuous re-estimation of task parameters (task difficulty) based on trial-by-trial feedback, which explains post-error accuracy improvement (supplementary sections S1.5-S1.6 and S2.5). In practice, learning about the task and outcome values is likely to be a more complex process, which may itself be altered in depression. Further to that, we only focused on the measures of working memory and attention, and did not tackle the known depression deficits in other domains such as learning and long-term memory [12,14,15,17]. These aspects of depression remain an important avenue for future investigations.

## CONCLUSION

Depression remains a prevalent and impactful condition which adversely affects cognitive function. In the current study we showed that deficits in working memory and attention at the DMS and RD tasks could be causally accounted for by the depression-related motivational factors – decreased belief of control and altered valuation of outcomes. Computational modelling made several predictions which could be directly tested experimentally by carefully evaluating cognitive performance measures against the predominant motivational symptoms. If the predictions are proven correct, cognitive performance at the tasks such as the DMS and RD could potentially serve as a basis for depression stratification and, with future research, for selection of optimal treatments.

## Data Availability

No specific datasets were analysed as part of the current study.

