## Supplementary Material for "Computational link between motivational factors and cognitive deficits in depression"

Aleks Stolicyn[12]
Liana Romaniuk[1]
Stephen M. Lawrie[1]
Peggy Seriès[2]

1. Division of Psychiatry
Centre for Clinical Brain Sciences, University of Edinburgh
Chancellor's Building, 49 Little France Crescent
Edinburgh BioQuarter, Edinburgh EH16 4SB, UK.

2. Institute for Adaptive and Neural Computation
School of Informatics, University of Edinburgh
10 Crichton Street, Edinburgh EH8 9AB, UK.

Corresponding author:

Aleks Stolicyn
Division of Psychiatry, University of Edinburgh
Chancellor's Building, 49 Little France Crescent
Edinburgh BioQuarter, Edinburgh EH16 4SB, UK


### S1. *Delayed Match to Sample Task Model Details*

#### S1.1 DMS Task General Model Description

In the current study we construct a very general formalisation of the DMS task which links performance to motivational factors, and which is applicable to different task implementations. We consider that each visual pattern in the task consists of $n$ distinct features, where each feature can take one of $k$ distinct values. In most DMS implementations, the features are colours and shapes of the separate quadrants of the pattern. At the selection stage, the participant is presented with $m$ patterns ($m = 4$ in CANTAB DMS). We define the proportion of features with the same values across all patterns (i.e. proportion of overlapping features) as $D$ ($D \in (0,1)$). $D$ can be directly interpreted as the difficulty of the task – the higher it is, the more features the participant has to memorise to perform correctly. The actual number of overlapping features is always whole, and hence can be defined as $d = round(D \cdot n)$. Due to limited cognitive resources, the participant only memorises $f$ features from the initial sample and attempts to identify the pattern at the selection stage based on these features ($f \in [0, n]$). Errors occur when the participant matches the $f$ memorised features to an incorrect (non-match) pattern because the match and non-match patterns happen to share these features (feature overlap).

Overall, DMS task performance in the model follows the following cycle:

1) Participant estimates the optimal number of features $f_{opt}$ to be memorised at the upcoming DMS trial so as to maximise utility $U$, drawing on their knowledge of the task ($n, k, m$ parameters and the correct response probability function $P_c$), their current estimate of the task difficulty ($D_{est}$), their belief of task controllability ($\alpha_c$), and their valuation of feedback and cognitive effort ($V_c, V_e, \eta_c$) (sections S1.2-1.4

below);

2) Participant deploys the cognitive resources to memorise the $f_{opt}$ features from the original sample and executes the trial by finding the pattern with matching features at the selection stage;

3) Participant receives trial feedback $F \in \{0, 1\}$, which they use to update their estimate of the task difficulty $D_{est}$ with feedback integration rate $\lambda_c$. The updated $D_{est}$ is then used in the subsequent trial. Feedback integration gives rise to the post-error improvement in accuracy (PIA) effect and the feedback-related negativity (FRN) signal (sections S1.5-1.7 below).

**S1.2 DMS Correct Performance Probability Function**

We here consider the version of the task where $d$ features are forced to overlap across all patterns and the remaining $n - d$ features are free to vary with the constraint that the original sample is unique at the selection stage (i.e. it has at least one feature that differentiates it from all other patterns). Probability of correct performance at a single DMS trial $P_c$ depends on the task parameters $n, k, m, d$ and the number of features $f$ that the participant memorises. To derive this probability function, we first define the probability of erroneously matching a single pattern at the selection stage.

We define $l$ as the number of features among those $f$ memorised by the participant that were not forced to overlap, $l \leq f$. Drawing on the values $n$, $k$ and $d$, the probability of any random non-match pattern being erroneously identified as a match pattern (an *error-match*) for any given value of $l$ is given by:

$$P_e(l) = \frac{k^{(n - d - l)} - 1}{k^{(n - d)} - 1} \tag{S1}$$

In the above equation, $k^{(n-d)}-1$ (the denominator) is the number of all possible non-matching patterns given one particular matching sample, and $k^{(n-d-l)}-1$ (the numerator) is the total number of patterns that the participant would erroneously match based on the $l$ memorised features.

Based on Equation S1 above, we have analytically derived the function of probability of correct performance at a single DMS trial, dependent on the number of features $f$ memorised by the participant. This probability is given by:

$$P_c(f) = 1 - \sum_{l=l_{min}}^{l_{max}} \left( \frac{C_f^l \cdot C_{n-f}^{n-d-l}}{C_n^d} \cdot \sum_{i=1}^{m-1} \left( \frac{i}{i+1} \cdot P_e^i(l) \cdot (1 - P_e(l))^{(m-1-i)} \cdot C_{m-1}^i \right) \right) \quad \text{(S2)}$$

In the above equation $l_{min} = max(0, f - d)$ is the minimum number of memorised features which become useful at the selection stage (i.e. are not forced to overlap between all patterns), and $l_{max} = min(f, n - d)$ is the maximum number of such spared memorised features. We here use $C_n^k$ as the notation for binomial coefficient, $C_n^k = \binom{n}{k} = \frac{n!}{k!(n-k)!}$.

The break-down of Equation S2 above is following:

1) The term $P_e^i(l) \cdot (1 - P_e(l))^{(m-1-i)}$ represents the probability of $i$ patterns (of $m$ in total) at the selection stage being *error-matches*, and respectively $m-1-i$ patterns being those that would be correctly identified as non-matches, based on the $l$ features. The term $C_{m-1}^i$ represents the number of pattern combination possibilities at the selection stage where $i$ patterns would be error-matches. Finally, the term $\frac{i}{i+1}$ represents the probability that the participant selects an error-match as opposed to the correct sample among all $i+1$ that they would identify as matching based on their $l$ memorised features (participant selects randomly among $i+1$ matching pattern candidates). There are at least two patterns at the selection stage, and hence $i$ can take

any value between 1 and $m-1$. The probability of an error at the selection stage for any given value $l$ is therefore given by the sum of error probabilities across all possible values of $i$: $\sum_{i=1}^{m-1}\left(\frac{i}{i+1} \cdot P_e^i(l) \cdot (1 - P_e(l))^{(m-1-i)} \cdot C_{m-1}^i\right)$.

2) The term $\frac{C_f^l \cdot C_{n-f}^{n-d-l}}{C_n^d}$ represents the probability that $l$ out of $f$ memorised features are not forced to overlap across all patterns. Here $C_n^d$ is the number of all possible configurations of $d$ out of $n$ overlapping features across all patterns; $C_f^l$ is the number of possible configurations of $l$ out of $f$ memorised features being not forced to overlap; and $C_{n-f}^{n-d-l}$ is the number of possible configurations of $n-d-l$ features being not forced to overlap but also not memorised by the participant, among all $n-f$ features not memorised by the participant.

3) The value of $l$ can vary randomly between $l_{min}$ and $l_{max}$ (please see above for $l_{min}$ and $l_{max}$ definitions), and hence the overall probability of making an error at a single trial is the sum of probabilities of making an error for all possible values of $l$, each weighted by the respective probability of the value of $l$ occurring (combination of the equations in points 1 and 2 above):

$$P_{et}(f) = \sum_{l=l_{min}}^{l_{max}}\left( \underbrace{\frac{C_f^l \cdot C_{n-f}^{n-d-l}}{C_n^d}}_{\text{probability of } l \text{ memorised features not overlapping}} \cdot \underbrace{\sum_{i=1}^{m-1}\left(\frac{i}{i+1} \cdot P_e^i(l) \cdot (1 - P_e(l))^{(m-1-i)} \cdot C_{m-1}^i\right)}_{\text{probability of an error at trial with } l \text{ memorised non-overlapping features}} \right)$$

Probability of correct response is then simply given by $P_c(f) = 1 - P_{et}(f)$, which ultimately leads to the Equation S2 above.

#### S1.3 DMS Control Belief Integration

We consider that when estimating the probability of success, the brain takes into account belief of task controllability $\alpha_c$, which is defined as the probability of contingency between actions and outcomes, consistently with the second notion of behavioural control in Huys & Dayan (2009) [1]:

$$P_{cw}(f) = P_c(f)\cdot\alpha_c + \frac{1}{m}\cdot(1 - \alpha_c) \quad \text{(S3)}$$

Note that in the above equation $P_c(f)$ is the *objective* probability of correct performance, assuming that the task is perfectly controllable. $\frac{1}{m}$, on the other hand, is the objective probability of correct performance when a *random* (instead of the initial) pattern at the selection stage is correct, i.e. there is no contingency between the number of memorised features (the action) and the feedback (the outcome), meaning that the task is not controllable. High controllability belief ($\alpha_c$) makes the estimated outcome probability dependent on the memorised features $f$, while low $\alpha_c$ biases the estimated probability to be similar across different values of $f$.

**S1.4 DMS Utility Optimisation**

We consider that the brain assigns values to correct and error feedback, which we respectively define as $V_c$ and $V_e$. Further, we consider the number of memorised features $f$ to be directly proportional to the amount of cognitive resources deployed, where memorising each feature is effortful and has a cost of $\eta_c$ [2,3]. Utility of memorising $f$ features at a single trial can be defined as the value expected to be gained after expending cognitive costs (mental effort):

$$U(f) = \underbrace{V_c\cdot P_{cw}(f) + V_e\cdot(1 - P_{cw}(f))}_{\text{expected outcome value}} - \underbrace{\eta_c\cdot f}_{\text{cognitive costs}} \quad \text{(S4)}$$

The definition of utility above is consistent with the EVC theory [4,5]. Optimal cognitive resource allocation at the DMS task requires identifying the number of features $f_{opt}$ which maximises utility and thus offers the best trade-off between the expended cognitive resources and the received reward:

$$f_{opt} = \underset{f}{argmax}(U(f)) \quad \text{(S5)}$$

**S1.5 DMS Trial Feedback Integration**

We consider that most DMS task parameters are either easily observable ( $m$ ) or can be estimated during the course of several trials ( $n$ , $k$ ). The main parameter which is not known and not easily observable for the participant is the task difficulty $D$ (i.e. the proportion of overlapping features). We suggest that the brain uses trial-by-trial feedback to continuously re-estimate this parameter during task performance. At each trial of the DMS task, participant is modelled to use their current difficulty estimate $D_{est}$ to compute $f_{opt}$ according to Equations S2-S5 above. The brain then deploys cognitive resources for memorising $f_{opt}$ features and the participant executes the trial. Based on the received trial feedback $F$ ( $F \in \{0, 1\}$ ), the participant updates their estimated probability of correct performance for $f_{opt}$ memorised features, with feedback integration rate $\lambda_c$ :

$$P'_{f_{opt}} = P_c(f_{opt}) \cdot (1 - \lambda_c) + \lambda_c \cdot F \quad \text{(S6)}$$

With the updated correct performance probability $P'_{f_{opt}}$ for $f_{opt}$ features, the difficulty parameter $D_{est}$ is re-estimated so that the updated probability function in Equation S2 fits the constraint of $P_c(f_{opt}) = P'_{f_{opt}}$ , given that all parameters apart from $D_{est}$ (i.e. $n, k, m$ ) are known and fixed. The re-estimated $D_{est}$ parameter is then used for probability computations (Equations S2-S5) at the following trial. In the simulations, we bounded the difficulty estimate to incorporate a rudimentary assumption that the task can be neither trivial, nor impossible $D_{est} \in (0.2, 0.8)$ .

**S1.6 DMS Post-error Improvement in Accuracy Effect**

The PIA effect in the model naturally arises due to continuous trial-by-trial re-estimation of the task difficulty (section S1.5 above). Success (correct) feedback ( $F = 1$ ) decreases the task difficulty estimate $D_{est}$ , while error feedback ( $F = 0$ ) increases it. When the task difficulty estimate is increased, optimal utility is computed for a higher number of memorised features (Figure 3B in the main text). This in turn leads to an increased probability of correct performance, and thus increased accuracy specifically after errors – the PIA effect. We compute the PIA effect in the following way in the simulations [6,7]:

$$PIA = P(correct \mid error\,previous) - P(correct \mid correct\,previous) \tag{S7}$$

**S1.7 DMS Feedback-Related Negativity Model**

To compute the FRN signal we draw on the grounded theoretical propositions that this signal reflects valued prediction error [8–11]. The FRN in our model is thus computed as the difference between the expected and the actual outcome values:

$$FRN = \underbrace{P_{cw}(f_{opt})\cdot V_c + (1 - P_{cw}(f_{opt}))\cdot V_e}_{expected\ outcome\ value} - \underbrace{(F\cdot V_c + (1 - F)\cdot V_e)}_{actual\ outcome\ value} \tag{S8}$$

**S1.8 DMS Model Parameter Specification**

To summarise, the DMS task in our model is specified by 4 parameters for task stimuli and difficulty ( $n,\ k,\ m,\ D$ ), while the participant is defined by another 5 control belief and valuation parameters ( $\alpha_c,\ V_c,\ V_e,\ \eta_c,\ \lambda_c$ ) (Table S1).

Within the simulations, $n$ , $k$ and $m$ parameters were specified to approximate the CANTAB version of the DMS task, and difficulty $D$ (proportion of overlapping features) was set fixed to $\frac{1}{2}$ . Control belief parameter ( $\alpha_c$ ) in healthy participants was set close to 1 to reflect a belief that the task is highly controllable (note that $\alpha_c \in (0,1)$ ). Correct and error feedback values were respectively set to canonical values of 1 and -1. We then specified the cognitive resource cost ( $\eta_c$ ) parameter so as to achieve an approximate 90% accuracy, which resembles typical performance of control participants at the DMS task [12,13]. Feedback integration rate parameter ( $\lambda_c$ ) regulates the PIA effect (Equations S6 and S7), and was specified to achieve an approximate 5% accuracy improvement after errors. The values of the DMS model parameters specified for healthy control participants are presented in Table S3.

#### S1.9 DMS Depression Modelling Details

Please see the methods and results sections of the main text for the details of the depression modelling constraints and the summary of the DMS depression simulation results. The two compared depression models were constrained to produce an approximate 10% decrease in accuracy, which corresponded to a 79% decrease in reward value ( $V_c$ ) and a 40% decrease in control belief ( $\alpha_c$ ). The exact parameter values are presented in Table S4.

### S2. *Eriksen Flanker Task Model Details*

#### S2.1 EF Task General Model Description

We applied the drift-diffusion framework of Dillon et al. (2015) [14], which was adapted from Noorani & Carpenter (2013) [15], to model reaction times and error rates at the EF task. Participant response at each trial is defined by three drift-diffusion processes: (1) prepotent drift which accumulates evidence from the flankers; (2) inhibitory control drift which prevents prepotent response; (3) executive control drift which accumulates evidence from the target (Figure 4 in the main text). Each drift process accumulates evidence until a predefined threshold, with a certain amount of noise (diffusion) to model variability in the perceptual system and the environment. At congruent trials, inhibitory and executive control processes are inactive and the response time is defined exclusively by the prepotent process. At incongruent trials, the prepotent drift process competes with the inhibitory control drift process. If the prepotent process wins (achieves the evidence threshold first), flanker-indicated incorrect response is executed, which results in an error. If the inhibitory control overruns the prepotent process, the erroneous response is prevented and the correct response is performed by the executive drift process. The inhibitory process exclusively controls the accuracy of performance at incongruent trials. Finally, the executive process is offset by a constant which reflects the time necessary to focus on the target symbol, and exclusively defines the response times at correct incongruent trials. Overall, the framework is specified by seven parameters: drift rates $v_{pre}$, $v_{inh}$ and $v_{exec}$, infinitesimal diffusion process variance $\sigma^2$, response threshold $a$, response motor delay $t_{motor}$, and executive control delay $t_{delay}$.

Decision (first-passage) times of the three drift processes are described by inverse Gaussian distributions (as for any Wiener processes):

$$\begin{aligned} t_{pre} &\sim IG\left(\frac{a}{v_{pre}}, \frac{a^2}{\sigma^2}\right) \\ t_{inh} &\sim IG\left(\frac{a}{v_{inh}}, \frac{a^2}{\sigma^2}\right) \\ t_{exec} &\sim IG\left(\frac{a}{v_{exec}}, \frac{a^2}{\sigma^2}\right) \end{aligned} \tag{S9}$$

Response times in the model are defined mainly by the first-passage times (Equation S9 above). For congruent trials and error incongruent trials, response time depends mainly on the prepotent process. For correct incongruent trials, response time depends on the executive process:

$$t_{response} = \begin{cases} t_{pre} + t_{motor} & \text{if } t_{pre} > t_{inh} \\ t_{exec} + t_{delay} + t_{motor} & \text{otherwise} \end{cases} \tag{S10}$$

We consider the inhibitory and executive drift processes to be effortful due to their relation to goal-driven cognitive control, and thus to incur cognitive costs proportional to the drift rates. In our account the prepotent drift rate ($v_{pre}$) is fixed, while the inhibitory and executive drift rates ( $v_{inh}$ and $v_{exec}$ ) are optimised to achieve the best reward-effort trade-off at incongruent trials.

EF task performance in the model follows the following cycle:

1) Participant estimates the optimal drift rates $v_{inh}^{opt}$ and $v_{exec}^{opt}$ so as to maximise respectively the single-trial utility $U$ and utility rate $UR$ , drawing on their knowledge of the task (estimated correct response probability function $P_c$ ), their current estimate of the task difficulty ( $D_{est}$ ), their belief of task controllability ( $\alpha_c$ ), and their valuation of correct and error responses and cognitive effort ( $V_c$, $V_e$, $\eta_c$ ) (sections S2.2-2.4 below);

2) Participant deploys the cognitive resources to enable drift rates $v_{inh}^{opt}$ and $v_{exec}^{opt}$ and executes the trial with these drift rates;

3) Participant receives either explicit or implicit trial feedback $F \in \{0,\ 1\}$ , which they use to update their estimate of the task difficulty $D_{est}$ with feedback integration rate $\lambda_c$ . The updated $D_{est}$ is then used in the subsequent trial. Feedback integration gives rise to the post-error improvement in accuracy (PIA) effect and the error-related negativity (FRN) signal (sections S2.5-2.6 below).

**S2.2 EF Correct Performance Probability Function**

Probability of an execution time ( $t$ ) of each of the three drift processes with a given drift rate $v$ is defined by the inverse Gaussian probability density function:

$$P_t(t,v) = \sqrt{\frac{a^2/\sigma^2}{2\pi t^3}}\ \exp\left(\frac{-\frac{a^2}{\sigma^2}\left(t - \frac{a}{v}\right)^2}{2\left(\frac{a}{v}\right)^2 t}\right) \tag{S11}$$

Probability of an error at an incongruent trial is defined by the probability of the prepotent drift process achieving the evidence threshold faster than the inhibitory drift process (i.e. probability of the prepotent process time being lower than the inhibitory process time):

$$P_e(v_{inh}) = \int_{t_i=0}^{\infty} P_t(t_i, v_{inh}) \int_{t_p=0}^{t_i} P_t(t_p, v_{pre}) \; dt_p dt_i \qquad \text{(S12)}$$

When the proportion of incongruent trials is defined by $D$ ( $D \in (0,1)$ , interpretable as difficulty of the task), probability of correct performance at a single EF trial can be defined as follows:

$$P_c(v_{inh}) = 1 - D \cdot P_e(v_{inh}) \qquad \text{(S13)}$$

Although the $D$ parameter in the task is actually fixed, it is not known to the participant and is modelled to be continuously re-estimated in the brain as $D_{est}$ .

**S2.3 EF Control Belief Integration**

Similarly to the DMS task (section S1.3 above), we consider that the brain takes into account its belief of task controllability $\alpha_c$ ( $\alpha_c \in (0,1)$ ) when estimating the probability of correct performance:

$$P_{cw}(v_{inh}) = P_c(v_{inh}) \cdot \alpha_c + \frac{1}{2} \cdot (1 - \alpha_c) \qquad \text{(S14)}$$

Note that $\frac{1}{2}$ above represents the probability of correct performance when the task is uncontrollable and a random (instead of target-related) response is correct. As can be noted in Equation S14, high controllability belief ( $\alpha_c$ ) makes the estimated outcome probability dependent on the deployed inhibitory drift rate $v_{inh}$ , while low $\alpha_c$ biases the estimated probability towards $\frac{1}{2}$ independent of $v_{inh}$ .

**S2.4 EF Utility Optimisation**

Similarly to the DMS task (section S1.4 above), utility of deploying a particular inhibitory drift rate $v_{inh}$ at a single trial can be defined as the value expected after expending the cognitive costs (mental effort) to support the drift rate:

$$U(v_{inh}) = \underbrace{V_c \cdot P_{cw}(v_{inh}) + V_e \cdot (1 - P_{cw}(v_{inh}))}_{\text{expected outcome value}} - \underbrace{\eta_c \cdot v_{inh}}_{\text{cognitive costs}} \tag{S15}$$

To obtain the best reward-effort trade-off, inhibitory drift rate is identified (optimised) so as to maximise utility, similarly to the number of features at the DMS task (Equation S5):

$$v_{inh}^{opt} = \underset{v \in (v_{min}, v_{max})}{argmax} (U(v)) \tag{S16}$$

Note that $v_{inh}^{opt}$ is continuous and not bounded by other model parameters. In our model, we specify $v_{max}$ as the lowest inhibitory drift rate which results in an error probability approximately lower than 1% when all other parameters are specified. For simplicity, we specify minimal inhibitory drift rate as a fraction of the maximal inhibitory drift rate $v_{min} = \frac{1}{5} \cdot v_{max}$.

Executive drift process exclusively controls reaction times at correct incongruent trials, and thus affects the rate of reward acquisition or *utility rate*. In our model we approximate *utility rate* as the efficiency of the executive process itself, or the utility acquired per second of this process:

$$UR(v_{exec}) = \frac{U(v_{inh}^{opt}) - \eta_c \cdot v_{exec}}{\frac{a}{v_{exec}}} \tag{S17}$$

In the above equation $\frac{a}{v_{exec}}$ is the expected (mean) first-passage time of the executive drift process, $\eta_c \cdot v_{exec}$ defines the cognitive cost of the process and $U(v_{inh}^{opt})$ defines the estimated optimal utility of a single trial (Equations S15 and S16). Optimal executive drift rate can then be defined as the one maximising utility rate:

$$v_{exec}^{opt} = \underset{v \in (v_{min}, v_{max})}{argmax} \left( UR(v) \right) \tag{S18}$$

#### S2.5 EF Trial Feedback Integration and PIA Effect

Similarly to the DMS task, after computing optimal drift rates $v_{inh}^{opt}$ and $v_{exec}^{opt}$ and executing a single trial, the participant receives either explicit or implicit feedback $F \in \{0, 1\}$, which is used to update the estimate of task difficulty $D_{est}$. To update the difficulty estimate, first the estimate of correct performance probability for the current optimal inhibitory drift rate ( $v_{inh}^{opt}$ ) is re-calculated using the feedback value $F$ and the feedback integration rate $\lambda_c$ :

$$P'_{v_{inh}^{opt}} = P_c\left(v_{inh}^{opt}\right) \cdot \left(1 - \lambda_c\right) + \lambda_c \cdot F \tag{S19}$$

Based on $P'_{v_{inh}^{opt}}$ above, new $D_{est}$ is identified so that the probability function in Equation S13 fits the constraint of $P_c\left(v_{inh}^{opt}\right) = P'_{v_{inh}^{opt}}$. The re-estimated $D_{est}$ parameter is then used for probability computations at the following trial. Similarly to the DMS task model, we bounded the difficulty estimate parameter to $D_{est} \in (0.2, 0.8)$. PIA effect at the EF task arises for the same reason as at the DMS task (section S1.6 above), and is computed in the similar way (Equation S7).

#### S2.6 EF Error-Related Negativity Model

Computation of the ERN signal magnitude in the EF task model follows the same principles as in the DMS task model (section S1.7 above), with the ERN estimated as the difference between the expected and actual outcome values:

$$ERN = \underbrace{P_{cw}\left(v_{inh}^{opt}\right) \cdot V_c + \left(1 - P_{cw}\left(v_{inh}^{opt}\right)\right) \cdot V_e}_{\text{expected outcome value}} - \underbrace{\left(F \cdot V_c + (1 - F) \cdot V_e\right)}_{\text{actual outcome value}} \tag{S20}$$

**S2.7 EF Model Parameter Specification**

To summarise, the EF model in our account is parametrised by the single task difficulty parameter $D$, 5 drift-diffusion framework parameters ($t_{delay}$, $t_{motor}$, $v_{pre}$, $a$, $\sigma^2$), and 5 parameters which define the participant control belief and performance valuation ($\alpha_c$, $V_c$, $V_e$, $\eta_c$, $\lambda_c$) (Table S2). Drift rates $v_{inh}$ and $v_{exec}$ are optimised at each trial based on the other parameters and the difficulty estimate $D_{est}$.

EF model parameters were specified as follows. The executive delay ($t_{delay}$), motor delay ($t_{motor}$), prepotent drift rate ($v_{pre}$) and response threshold of the drift processes ($a$) were taken to closely approximate the values for control participants in the original account of the model in Dillon et al. (2015) [14]. Infinitesimal variance of the drift processes ($\sigma^2$) was not reported in this account and was computed to replicate the incongruent trial accuracy of 76% when the inhibitory drift rate is constrained to $v_{inh} = 9.76$ – these values were reported for control participants. Control belief parameter ($\alpha_c$) was set to be the same as in the DMS model. Correct performance value ($V_c$) was specified to the canonical value of 1. We then specified $V_e$ (error value) and $\eta_c$ (cognitive cost) parameters so as to achieve optimal drift rates $v_{inh}^{opt} = 9.7$ and $v_{exec}^{opt} = 10.4$ when the difficulty estimate is set to $D_{est} = 0.34$ (Equations S12-S18). These three constraints correspond to the control participant performance characteristics from Dillon et al. (2015) [14]. Error value specified in this way was $V_e = -0.2$, which contrasts with $V_e = -1$ in the DMS task model, while the reward value was similar in both cases ($V_c = 1$). This could potentially be explained by the absence of explicit feedback in Dillon et al. (2015), which likely decreased the error value. Finally, we specified the feedback integration rate parameter ($\lambda_c$) to approximate a 5% PIA effect, similarly to the DMS model. The resulting set of EF model parameters for healthy controls can be found in Table S5.

#### S2.8 EF Depression Modelling Details

Please see the methods and results sections of the main text for the details of the depression modelling constraints and the summary of the EF depression simulation results. The exact parameter values of the five investigated depression models are presented in Table S6 below. Exact effects of the five depression models over performance measures at the EF task are presented in Table S7.

**Table S1** Delayed Match to Sample task model parameters

| DMS Model Parameter | Description |
|---|---|
| $n$ | Number of features in a single stimulus pattern |
| $k$ | Number of values each stimulus feature can take |
| $m$ | Number of pattern stimuli at the selection stage |
| $D$ | Task difficulty – proportion of overlapping features between all patterns at the selection stage |
| $\alpha_c$ | Belief of controllability of the task (control belief) |
| $V_c$ | Value of correct outcome (feedback) at a single trial |
| $V_e$ | Value of error outcome (feedback) at a single trial |
| $\eta_c$ | Cognitive cost of memorising a single feature of the stimulus |
| $\lambda_c$ | Feedback integration rate for updating difficulty estimate |

**Table S2** Eriksen Flanker task model parameters

| EF Model Parameter | Description |
|---|---|
| $D$ | Task difficulty – proportion of incongruent trials |
| $v_{pre}$ | Prepotent (habitual response) process drift rate |
| $\sigma^2$ | Infinitesimal diffusion process variance |
| $a$ | Drift process response threshold |
| $t_{motor}$ | Response motor delay |
| $t_{delay}$ | Executive control process delay |
| $\alpha_c$ | Belief of controllability of the task (control belief) |
| $V_c$ | Value of correct outcome (feedback) at a single trial |
| $V_e$ | Value of error outcome (feedback) at a single trial |
| $\eta_c$ | Cognitive cost scaling factor of the drift rate |
| $\lambda_c$ | Feedback integration rate for updating difficulty estimate |

**Table S3** Delayed Match to Sample model parameter values for healthy controls

| **Parameter** | $n$ | $k$ | $m$ | $D$ | $\alpha_c$ | $V_c$ | $V_e$ | $\eta_c$ | $\lambda_c$ |
|---|---|---|---|---|---|---|---|---|---|
| ***Value*** | 8 | 4 | 4 | 0.5 | 0.975 | 1 | - 1 | 0.12 | 0.125 |

**Table S4** Delayed Match to Sample depression model parameter values

| | $\alpha_c$ (control belief) | $V_c$ (reward value) |
|---|---|---|
| Healthy Control | 0.975 | 1 |
| Reward Sensitivity Deficit | 0.975 | *0.233* (↓79%) |
| Control Belief Deficit | *0.6* (↓40%) | 1 |

*Note*: Percentage numbers in brackets indicate deficit-related change relative to healthy controls.

**Table S5** Eriksen Flanker model parameter values for healthy controls

| **Parameter** | $v_{pre}$ | $a$ | $\sigma$ | $t_{delay}$ | $t_{motor}$ | $\alpha_c$ | $V_c$ | $V_e$ | $\eta_c$ | $\lambda_c$ |
|---|---|---|---|---|---|---|---|---|---|---|
| ***Value*** | 7 | 1.1 | 0.975 | 130 ms | 210 ms | 0.975 | 1 | - 0.2 | 0.029 | 0.07 |

**Table S6** Eriksen Flanker depression model parameter values

| | $\alpha_c$ (control belief) | $V_c$ (reward value) | $V_e$ (error value) |
|---|---|---|---|
| Healthy Control | 0.975 | 1 | - 0.2 |
| Control Belief Deficit | *0.725* | 1 | - 0.2 |
| Reward Sensitivity Deficit | 0.975 | *0.855* | - 0.2 |
| Negative Value Deficit | 0.975 | 1 | *- 1.75* |
| Control Belief & Negative Value Deficits | *0.725* | 1 | *- 0.6* |
| Reward Sensitivity & Negative Value Deficits | 0.975 | *0.855* | *- 0.35* |

**Table S7** Depression model effects over EF task performance

| | *Incongruent Correct RT* | *Incongruent Error RT* | *Incongruent Correct Rate* | *PIA* | *ERN* |
|---|---|---|---|---|---|
| Control Belief Deficit | + 31.3 ms * | + 7.1 ms * | - 9.8% * | - 0.04% | - 0.16 * |
| Reward Sensitivity Deficit | + 32.7 ms * | + 3.1 ms * | - 3.6% * | + 0.18% | - 0.15 * |
| Negative Value Deficit | + 15.9 ms * | - 21.1 ms * | + 16.4% * | - 1.62% * | + 1.57 * |
| Control Belief & Negative Value Deficits | + 59.1 ms * | + 0.4 ms | - 0.04% | - 0.04% | + 0.1955 * |
| Reward Sensitivity & Negative Value Deficits | + 36 ms * | - 0.7 ms | + 0.13% | - 0.14% | + 0.0057 * |

* = $p < 0.00001$
